# Accelerating Functional Endpoints in Geographic Atrophy Trials via Morphology-Based Perimetry Grids

**DOI:** 10.64898/2026.08.10.26360122

**Authors:** Giovanni Ometto, Giovanni Montesano, Alison Binns, Christiana Dinah, David P. Crabb

## Abstract

**Purpose:** To evaluate whether a Geographic Atrophy Morphology-based Mapping Algorithm (GAMMA) grid, informed by geographic atrophy (GA) lesion morphology, can accelerate functional progression detection compared with a conventional 10-2 grid and a dense grid (129 locations). This work is motivated by emerging regulatory expectations requiring at least five locations to worsen by ≥7 dB from baseline.

**Methods:** Binary atrophy masks from six autofluorescence images were used to simulate GA expansion over 3 years at 3-month intervals using a stochastic perimeter-growth model with a fixed preferential expansion direction (P_dir_). For each image, 32 independent growth histories and 32 microperimetric test realisations per history were generated. For each grid (10-2, Dense, and GAMMA), 5-point clusters were selected outside the baseline GA lesion along three directions (0°, 30°, 120°) away from P_dir_, simulating full, partial, and no prior knowledge of P_dir_. Ground-truth sensitivities were <0 dB inside the GA lesion and normal outside, calculated using a published normative equation. Response variability was simulated following Henson et al. with baseline averaging. Detection time was the first visit at which all five selected locations showed ≥7 dB loss from baseline. Survival curves and median detection times (T50) were used to compare grid performance.

**Results:** The GAMMA grid achieved the earliest progression detection across all scenarios. Under full knowledge of the expansion direction, T50 was 1.0 year for GAMMA versus 1.25 and 1.5 years for Dense and 10-2, respectively. With partial knowledge, GAMMA’s T50 was 1.25 years versus 1.5 and 2.0 years for Dense and 10-2. Even under no knowledge, GAMMA detected progression earliest (T50 = 1.5 years), while Dense required 6 months longer and 10-2 nearly double the time (2.75 years).

**Conclusions:** The automatic GAMMA grid accelerates detection of localised functional progression compared with conventional and dense grids. Structure-informed grid optimisation may better align testing with likely expansion paths, potentially reducing follow-up duration and sample sizes in perimetry-based interventional trials.

## INTRODUCTION

Geographic atrophy (GA), the atrophic, late form of age-related macular degeneration (AMD), is a leading cause of irreversible central visual impairment in the UK, responsible for approximately one quarter of all certifications as sight-impaired in England and Wales(1,2). After decades without approved treatments, two complement inhibitors, pegcetacoplan and avacincaptad pegol, received FDA approval in 2023 based on anatomical endpoints, with further therapies in Phase III trials(3). However, a major bottleneck in GA trial design remains the poor correlation between anatomical and functional endpoints(4–8), contributing to lack of regulatory approval in many jurisdictions and poor uptake in regions with approval. Best corrected visual acuity (BCVA), the traditional functional endpoint, is poorly suited to GA because it is a measure of cone-mediated foveal spatial resolution (or the preferred retinal locus in foveal-involving GA) and GA characteristically spares the central fovea till late in the disease. BCVA has therefore failed to separate treated from sham arms in pivotal trials so far. This has driven a search for functional assessments sensitive to the localised, parafoveal involvement typical of GA progression. Microperimetry, which maps retinal sensitivity at spatially defined locations co-registered to the atrophic lesion, has emerged as one of the most promising candidates and the assessment regulators have pointed to as a benchmark for demonstrating functional efficacy. Specifically, regulatory expectations suggest a minimal clinically important difference of at least five pre-specified microperimetric locations to worsen by ≥7 dB from baseline, adapted from the standard automated perimetry benchmark used in glaucoma trials(9). This threshold sits comfortably above the -6 dB point-wise coefficient of repeatability now reported for microperimetry in GA cohorts(10,11). Critically, however, this criterion specifies how many locations must progress without specifying where they should lie or at what spatial density they should be sampled(12). The dominant 10-2 grid evenly distributes 68 test points across the central 10° visual field. However, a proportion inevitably fall outside the zone of fastest GA expansion or inside already-atrophic retina, and test-retest variability across multiple points increases the risk of false-positive results, undermining regulatory trust and requiring either longer trials or larger cohorts, increasing cost, patient burden, and time to treatment access(13). A wave of microperimetric innovation has addressed adjacent problems, such as patient-tailored iso-contour grids that concentrate resolution at the lesion border(14), defect-mapping strategies that track deep-scotoma extent(15) and machine-learning inference of dense sensitivity maps from structure(16). Nevertheless, the question of how to position a fixed, small number of test locations so that they capture progression fastest remains largely unaddressed and is the question the present work seeks to address.

The GAMMA (Geographic Atrophy Morphology-based Mapping Algorithm) testing strategy builds on a patented algorithm(17) that converts a structural retinal map, such as the GA lesion boundary on fundus autofluorescence (FAF), into a ranked list of candidate test locations, ordered from most to least informative for describing a lesion’s shape. In principle, each patient’s baseline FAF generates a ranked list across the central visual field, from which a fixed number of highest-ranking locations can be tested at each visit, producing a personalised sensitivity map directly comparable to the 10-2 in coverage and test count. In the present, post-hoc analysis, five GAMMA locations and five 10-2 locations, selected by the same criterion based on the direction of historical lesion expansion, were compared for their ability to meet the regulatory-aligned detection threshold, alongside a third, denser but morphology-agnostic 129- location grid.

The fastest-progressing GA boundary, identified from pre-baseline FAF timepoints, was used to identify the five closest candidate locations in each grid. FAF-derived boundary expansion rate is among the most reliable available biomarkers for predicting future lesion growth and expected structure-function change(5,18). We previously showed that GAMMA-derived grids substantially outperform the 10-2 grid in targeting structurally relevant locations(19). Here, we extend this work by applying a regulatory-aligned functional benchmark directly, testing the hypothesis that a GAMMA grid, built around a patient’s directions of fastest expansion, translates into earlier detection of the multi-location, cluster-based functional progression criterion than either the 10-2 grid or the denser alternative, and that this advantage persists even when the true direction of lesion expansion is only partially, or not at all, known in advance.

## METHODS

### Study images and lesion growth simulation

Binary atrophy masks were segmented from baseline FAF images of six eyes with GA and used as the starting lesion boundary for each simulation. GA expansion was simulated over three years at 3-month intervals (13 timepoints) on a fixed 768×768-pixel grid (pixel size 0.01157 mm, ∼8.9 mm field, matching Spectralis FAF imaging geometry) using a stochastic, perimeter-driven growth model. At each simulation step, boundary-adjacent pixels were candidates for new atrophy; the number of pixels converted was calibrated to a constant target radial expansion speed (0.3 mm/year) regardless of local boundary length, so that irregularly-shaped lesions did not spuriously expand faster than round ones purely by virtue of having a longer perimeter.

Each candidate pixel’s probability of being selected combined three multiplicative weights: a surface-tension term favouring pixels with more already-atrophic neighbours (promoting smooth, rounded expansion rather than fingering artefacts); a fixed foveal-sparing repulsor (exponential decay, 1.2 mm) that suppressed growth near the fovea; and the net effect of a set of local influence points, each an exponentially-decaying attractor (growth-promoting) or repulsor (growth-suppressing) combined multiplicatively across overlapping influences. For each eye, one fixed attractor (equivalent to a 3-fold local growth-rate boost at its centre, 1.0 mm decay, located 2.5 mm from the fovea) anchored a consistent, eye-specific preferential expansion direction (P_dir_) shared by every growth history for that eye, while 15 additional points, randomly positioned 1.0–3.5 mm from the fovea and independently, randomly assigned as attractors or repulsors (random strength magnitude, 0.15–0.7; 0.1–0.5 mm decay), varied between growth histories to introduce realistic local irregularity without altering the eye’s overall directional bias. At each step, candidate pixels were ranked by weight multiplied by an independent random draw and the top-ranked pixels converted to atrophy; thirty-two independent growth histories were generated per eye by resampling these 15 random points, producing simulated variation in expansion shape and rate while preserving each eye’s underlying P_dir_.

### Test grids

Three perimetric test grids were compared, all confined to the same overall central visual field area. The 10-2 grid comprised 68 fixed locations, identical in every eye. The Dense grid comprised 129 locations covering the same field at higher, uniform spatial resolution. The GAMMA grid was generated individually for each eye using a patented, informativeness-ranking algorithm(17) and, unlike the other two grids, was built from the anticipated future extent of the lesion rather than from its baseline shape alone. Specifically, each eye’s baseline mask was dilated isotropically (i.e., direction-agnostically, by simple uniform expansion rather than the directionally-biased growth model above) at the average expansion rate of 0.3 mm/year, generating a staircase map with equally-spaced intensity levels representing the lesion’s isotropic 3-year growth envelope; this map, not the raw baseline shape, was used as the algorithm’s input (**Fig 2**). Because this dilation was direction-blind, the resulting candidate grid had no privileged knowledge of the eye’s true preferential expansion direction (P_dir_), and the same candidate grid was reused across all 32 growth histories of a given eye. This forecast was converted into a synthetic sensitivity surface using the normative hill-of-vision equation above, and an information map was computed as its local curvature (the gradient of its gradient magnitude, after light smoothing), which peaks at the lesion margin, where sensitivity is changing most steeply, rather than within the deep scotoma or unaffected retina.

**Figure 1.**
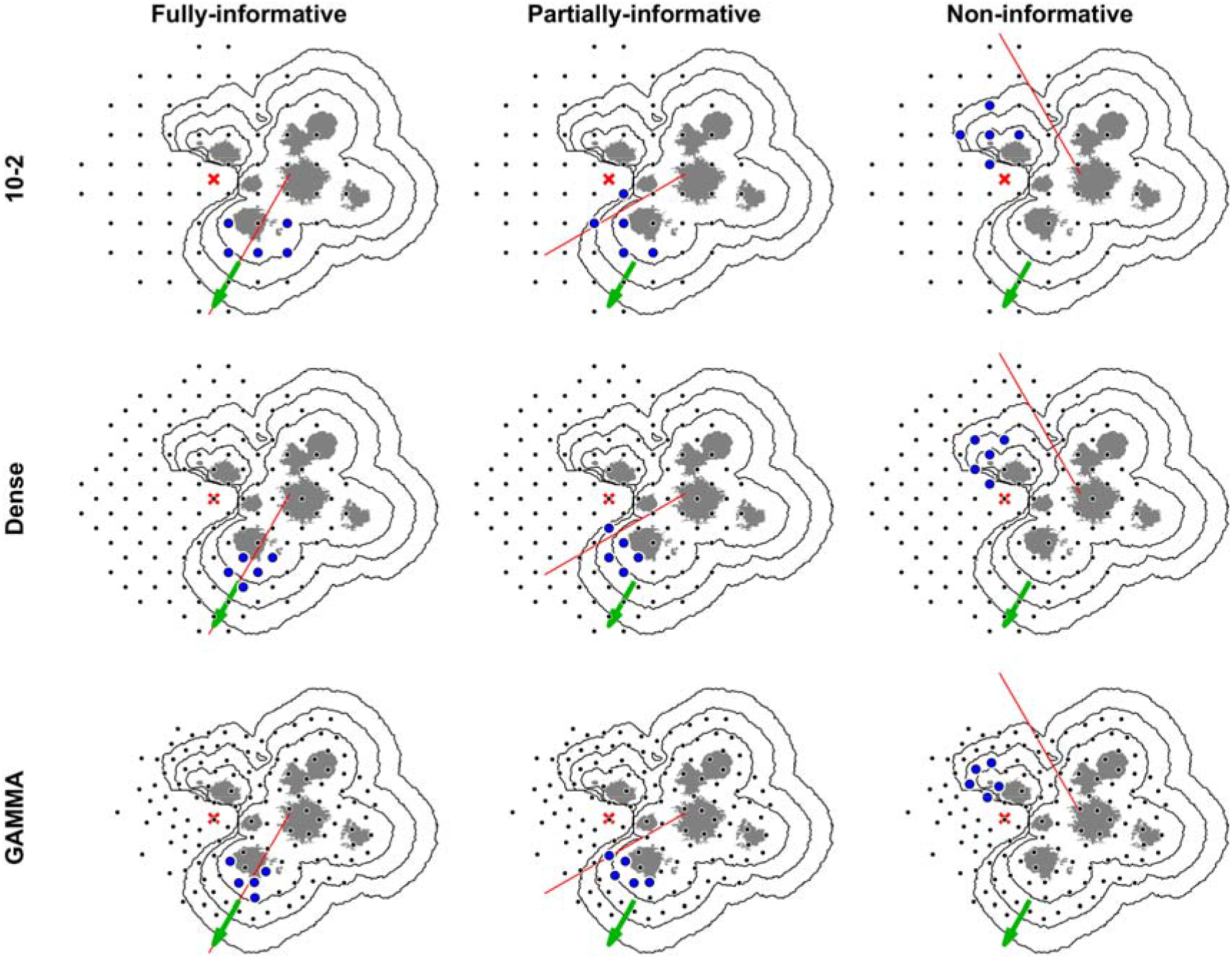
5-point cluster selection (blue dots) using 10-2, Dense, and GAMMA grids (rows) against simulated geographic atrophy expansion (black contours), under full, partial, and no prior knowledge of the true preferential growth direction (rows × columns as labelled: Fully-/Partially-/Non-informative). Green arrow: true preferential expansion direction (Pdir). Red line: assumed direction used for cluster selection. Red cross: fovea.

**Figure 2.**
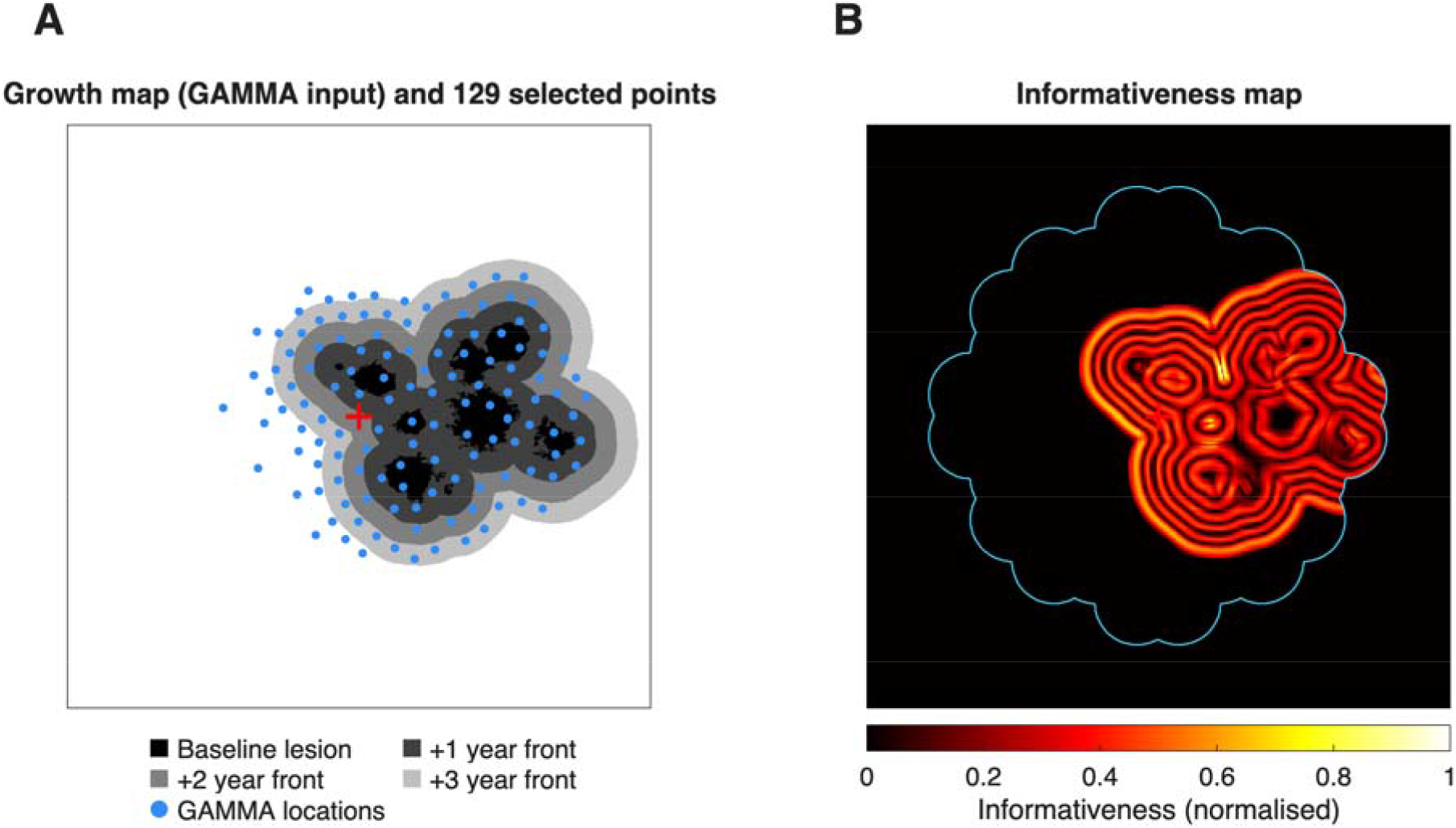
The GAMMA algorithm’s input and output, for the same eye shown in Figure 1. Fovea is displayed with a red cross. (A) The staircase, equally-spaced-intensity growth map, built by isotropically dilating the baseline lesion mask at the eye’s average radial expansion rate (0.3 mm/year) out to 1, 2 and 3 years; this direction-blind forecast is the GAMMA algorithm’s input. Blue dots: the 129 candidate locations the algorithm selected from this map. (B) The corresponding informativeness map computed by the algorithm from the growth map, from which the 129 locations are greedily selected. The light blue line represents the approximate area covered by a 10-2 grid.

This information map was further down-weighted at locations with high expected test-retest variability, using the same Henson-derived variability model applied to the perimetry simulation (weight 0.3 of the normalised, 6 dB-capped SD), so that candidate selection favoured informative and reliably-measurable locations over informative-but-noisy ones near the sensitivity floor. Candidate locations were then chosen greedily: at each iteration, the single most informative remaining pixel was added to the grid, after which a local exclusion zone was applied around it so that it could not be resampled. This exclusion radius adapted to the local variance of the information map (minimum 1.03°, approximating the average annual radial expansion distance used in the growth simulation at 0.3 mm/year), shrinking near highly informative, high-variance regions such as the lesion boundary to allow denser sampling, and expanding over flatter, less informative background to avoid redundant testing. This was repeated until 129 candidate locations were selected (matching the Dense grid), yielding a ranked, eye-specific candidate list ordered from most to least informative for describing that eye’s lesion shape and its anticipated expansion.

### Cluster selection

The three knowledge scenarios were applied at cluster-selection stage only, identically across all three grids, so that no grid had privileged access to the eye’s true expansion direction during grid generation itself (this applies in particular to GAMMA, whose candidate locations, as above, are generated from a direction-agnostic forecast). For each grid, a 5-point cluster was selected from among that grid’s existing candidate locations lying outside the baseline lesion, along one of three directions relative to P_dir_: 0° (aligned with the true expansion direction, “full knowledge”), 30° (“partial knowledge”), and 120° (“no knowledge”). These three conditions represent decreasing accuracy of a clinician’s a-priori estimate of the eye’s true direction of future expansion, as might be derived in practice from pre-baseline FAF timepoints.

### Simulated perimetry

At baseline and at each follow-up visit, sensitivity at the 5 cluster locations was simulated from a normative, age-corrected hill-of-vision equation (32.75 − 0.07 × age − 0.42 × eccentricity in degrees; age fixed at 60 years), combined with the underlying ground-truth lesion mask (sensitivity floored at the atrophic retina and Gaussian-smoothed at the lesion boundary to avoid a step discontinuity). Trial-to-trial response variability was added using a published normative model of perimetric variability(20), in which the standard deviation of the response increases exponentially as true sensitivity falls, capped at ±6 dB; baseline variability was reduced to reflect within-visit averaging over two tests. Thirty-two independent perimetric test realisations were simulated per growth history, giving 6 eyes × 32 growth histories × 32 test realisations = 6,144 simulated eye-histories per grid-direction combination (55,296 in total across 3 grids × 3 directions).

### Detection criterion and statistical analysis

An eye-history was scored as meeting the functional progression criterion at the first visit at which all five cluster locations showed a simulated sensitivity loss of ≥7 dB relative to that eye-history’s own simulated baseline. Time-to-detection was summarised with Kaplan-Meier-style survival curves (proportion not yet detected vs. time) for each grid × knowledge-level combination, with eye-histories not reaching criterion by 3 years treated as censored. Grids were compared on cumulative detection rate at 3 years and on the 25th/50th/75th-percentile detection times (T25/T50/T75) read from the survival curves.

## RESULTS

Across the six study eyes, the GAMMA grid achieved the highest cumulative detection rate and the shortest time to reach the five-location, ≥7 dB functional progression criterion in every knowledge scenario (**Table 1, Fig 3**).

**Table 1.**

| Grid | Knowledge of $P_{dir}$ | Detected by 3 yr (%) | T25 (yr) | T50 (yr) | T75 (yr) |
| --- | --- | --- | --- | --- | --- |
| 10-2 | Full | 84.6 | 1.25 | 1.50 | 2.75 |
| | Partial | 63.6 | 1.50 | 2.00 | $>3.00$ |
| | No | 59.2 | 2.25 | 2.75 | $>3.00$ |
| Dense | Full | 99.3 | 1.00 | 1.25 | 1.50 |
|  | Partial | 83.4 | 1.00 | 1.50 | 2.00 |
| | No | 63.5 | 1.75 | 2.00 | $>3.00$ |
| GAMMA | Full | 99.5 | 1.00 | 1.00 | 1.25 |
|  | Partial | 84.9 | 1.00 | 1.25 | 1.75 |
| | No | 69.5 | 1.25 | 1.50 | $>3.00$ |

Kaplan-Meier detection-time summary by grid and knowledge scenario ( $n = 6,144$ simulated eye-histories per cell: 6 eyes × 32 growth histories × 32 test realisations).

**Figure 3.**
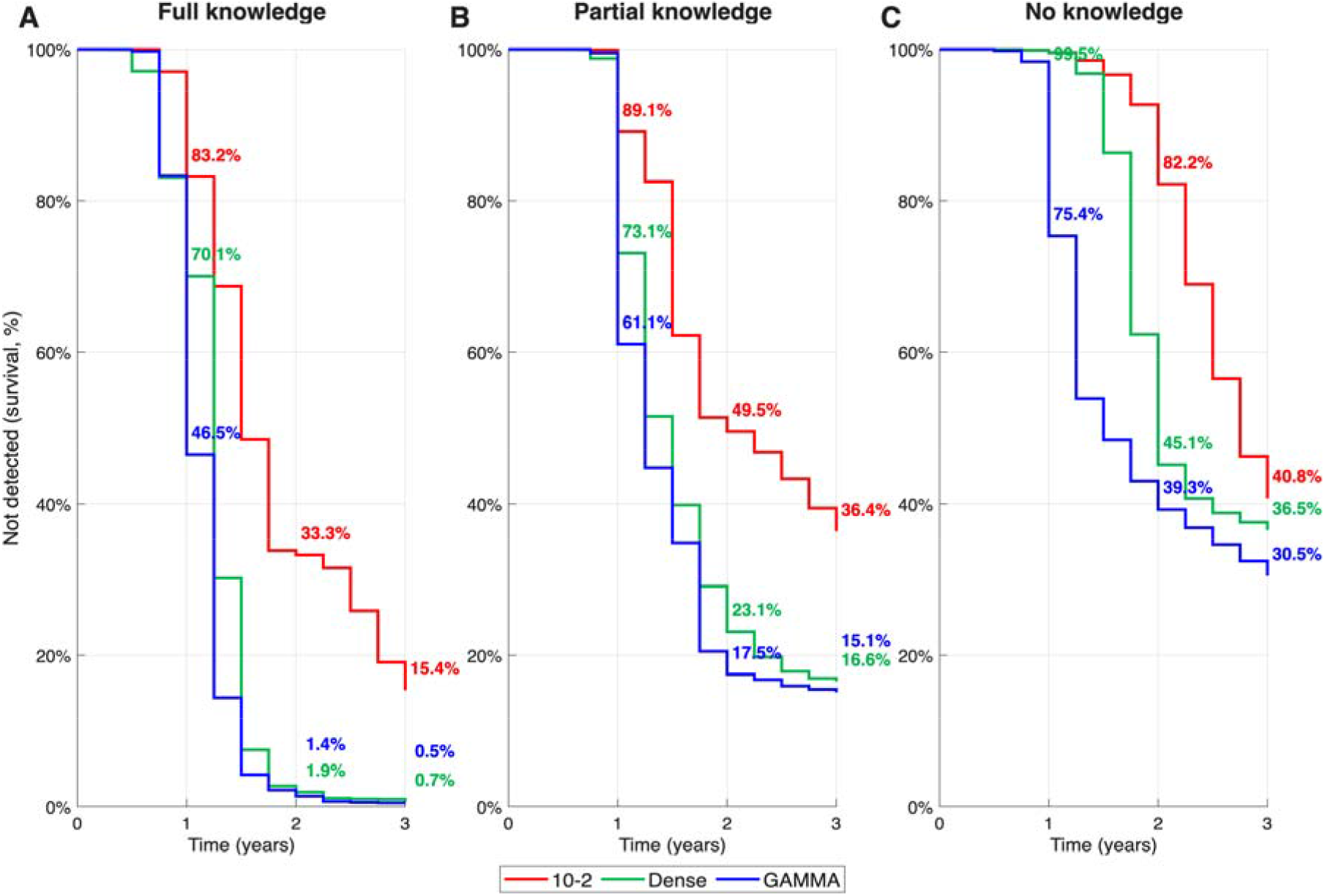
Kaplan-Meier-style survival curves (proportion of simulated eye-histories not yet meeting the ≥5-location, ≥7 dB detection criterion) over 3 years, stratified by knowledge of the true preferential expansion direction (A: full knowledge; B: partial knowledge; C: no knowledge) and grid (red = 10-2, green = Dense, blue = GAMMA). Annotated percentages mark survival (i.e., 100% minus cumulative detection rate) at each step change.

### Full knowledge of the expansion direction

By 1 year, 53.5% of GAMMA eye-histories had already met the detection criterion, compared with 29.9% for Dense and 16.8% for 10-2 (**Fig 3**A). Detection was essentially complete for both structurally-informed grids by 2 years (>98%), whereas 10-2 lagged throughout, reaching only 84.6% cumulative detection by 3 years. Median detection time (T50) was 1.00 year for GAMMA, 1.25 years for Dense, and 1.50 years for 10-2.

### Partial knowledge

The advantage of structurally-informed grids narrowed but persisted: at 1 year, 38.9% of GAMMA eye-histories were detected, versus 26.9% for Dense and 10.9% for 10- 2. T50 was 1.25 years for GAMMA, 1.50 years for Dense, and 2.00 years for 10-2; 3-year cumulative detection was 84.9%, 83.4%, and 63.6%, respectively.

### No knowledge

When the selected cluster direction was maximally uninformative about the eye’s true expansion path, GAMMA still detected progression earliest. The 75th-percentile detection time (T75) was not reached within the 3-year simulation horizon for any grid under no-knowledge conditions.

## DISCUSSION

In a simulation framework designed to mirror a cluster-based, five-location, ≥7 dB functional progression criterion, a lesion-morphology-informed test grid (GAMMA) detected functional progression earlier than either a conventional 10-2 grid or a denser, morphology-agnostic 129- location grid. This was observed across every level of prior knowledge of the eye’s true direction of fastest expansion: under full knowledge, median detection time fell from 1.5 years (10-2) to 1.0 year (GAMMA), a 33% reduction; under partial knowledge, from 2 to 1.25 years, a 37.5% reduction; under no knowledge, from 2.75 to 1.5 years, a 45% reduction. Notably, GAMMA’s T50 under this adverse scenario (1.50 years) equalled 10-2’s T50 under the most favourable scenario of full directional knowledge (also 1.50 years): GAMMA’s worst case matched 10-2’s best case. Under no knowledge, 10-2 required a T50 of 2.75 years, nearly double GAMMA’s. In this simulated setting, the 75th-percentile detection time (T75) was not reached within the 3- year horizon by any grid, suggesting that a minority of eye-histories will not meet the endpoint within a conventional 3-year trial window regardless of grid choice when the fastest direction of expansion is grossly misjudged. Because the sample size required for a time-to-event endpoint scales directly with the achieved event rate at a fixed follow-up duration, these differences imply that GAMMA-guided grids could support shorter and/or smaller functional-endpoint trials at equivalent statistical power. For example, assuming the magnitude of enhanced detection observed here is transferable to a clinical setting, and taking the partial-knowledge scenario as the realistic case, a trial designed to detect a hypothetical 30% reduction in hazard of functional progression (α = 0.10, two-sided; 80% power) would require approximately 435 participants (218 per arm) with a 24-month follow-up using the 10-2 grid, versus approximately 254 (127 per arm) using GAMMA, a 42% reduction in sample size for the same follow-up duration. Equivalently, for a fixed sample size of 435, GAMMA would reach the same statistical power after approximately 15 months of follow-up rather than 24.

GAMMA’s advantage did not depend on accurate prior knowledge of the expansion direction. Its performance in the worst-case (“no knowledge”) scenario matched 10-2’s performance in the best-case (“full knowledge”) scenario. In practice, the true future direction of GA expansion is never known with certainty at baseline and must be estimated from pre-baseline FAF timepoints, which are themselves an imperfect predictor. This robustness suggests that GAMMA’s benefit should be preserved when applied prospectively with realistic, imperfect estimates of expansion direction, rather than depending on an idealised best case that may not be achievable in a real trial population. GAMMA also sits within, rather than in competition with, a broader shift towards structurally-informed perimetry, and its distinguishing feature relative to these approaches is that point placement follows an explicit, automated, reproducible criterion rather than manual selection or fixed, uniform coverage. Patient-tailored microperimetry using iso-contour grids already represents a step in this direction: points are concentrated along the lesion border, but density is typically distributed evenly around its full circumference, since the approach has no basis for favouring one sector over another. GAMMA can be understood as a directional extension of this same iso-contour principle: rather than treating the border as uniform, it reallocates that same concentrated density towards the arc where expansion is anticipated, using a defined structural criterion (such as the expansion-direction information from prior FAF timepoints) to decide which portion of the contour merits denser sampling. The same criterion that guides GAMMA’s own point placement could, in principle, also be used to inform where defect-mapping strategies site their test locations, biasing coverage of deep- scotoma boundaries towards the segment most likely to advance next rather than mapping the full margin with uniform density. GAMMA is likewise complementary to machine-learning approaches that infer dense sensitivity maps directly from structural imaging: these generate a structure-derived prediction of function across the retina, whereas GAMMA converts a structural criterion into a small, discrete set of test-point coordinates, addressing the practical constraint that only a limited number of points can be tested in a clinical session.

These findings extend our earlier observation that GAMMA-derived grids more efficiently target locations of structural relevance than the 10-2 pattern(19), by showing that this structural advantage translates into an earlier functional, regulator-relevant endpoint. They are consistent with a wider literature identifying the disconnect between structural and functional endpoints as a central obstacle in GA trials(4–8), and with evidence that FAF-derived expansion kinetics are among the most reliable predictors of future structure-function change(5,18).

Several limitations should be considered. First, this is a simulation study: lesion growth, response variability, and the normative sensitivity model, while based on published data(20), are simplifications of true disease behaviour, which can include multifocal, coalescing, or non-monotonic expansion not captured by a single fixed preferential direction. Second, only six baseline lesion morphologies were used; although growth and test variability were resampled extensively (32 × 32 realisations per eye), the diversity of lesion shapes represented remains limited, and these findings should be confirmed in a larger, prospectively recruited cohort. Third, even the best-performing grid did not reach a 75% cumulative detection rate within 3 years under no-knowledge conditions, underscoring that structural targeting alone cannot fully overcome the difficulty of anticipating an unknown future growth path. In similar circumstances, some proportion of participants will remain below threshold within realistic follow-up windows regardless of grid choice.

In conclusion, structure-informed grid optimisation, targeting prespecified test clusters towards an eye’s most probable direction of GA expansion, accelerated detection of a regulatory-aligned functional progression criterion relative to both a conventional 10-2 grid and a denser, non-targeted alternative, with the benefit persisting under uncertain knowledge of the true expansion direction. These findings support prospective clinical evaluation of GAMMA-guided perimetry as a strategy to shorten follow-up duration and reduce sample-size requirements in GA interventional trials.

## Data Availability

All data produced in the present study are available upon reasonable request to the authors.

## DECLARATIONS

### Data availability

The MATLAB code used to generate the simulations and perform the analyses in this study is not publicly available.

